# Knowledge, attitudes, and practices regarding COVID-19 among university students and employees in Massachusetts, USA: a qualitative study

**DOI:** 10.1101/2022.07.05.22277273

**Authors:** Johanna Ravenhurst, Teah Snyder, Kate Wallace, Sheila Pennell, Sarah L. Goff, Andrew A. Lover

## Abstract

**Background:** At the time of this study, Massachusetts had recorded a total of 352,558 COVID-19 cases and 12,076 deaths. Few qualitative studies have been conducted that investigate the experiences of university students and employees during the COVID-19 pandemic. The objective of this study was to explore the knowledge, attitudes, and practices regarding COVID-19 in university affiliates to inform future COVID-19 policies.

**Methods:** Semi-structured focus groups and interviews were conducted via Zoom between December 14, 2020 and January 15, 2021. Twenty-two focus group participants included undergraduate students, graduate students and university employees who had not experienced isolation or quarantine during the Fall 2020 semester. Fourteen participants who had experienced quarantine or isolation were interviewed individually to protect confidentiality. Data were analyzed using Dedoose software via inductive thematic analysis, with reporting as per Consolidated Criteria for Reporting Qualitative Studies (COREQ) guidelines.

**Results:** Five major themes emerged from these data: COVID-19 knowledge, stress and coping, trust, decision-making, and institutional feedback. Misinformation regarding COVID-19 was common, compounding high levels of stress reported by many participants. Reported direct sources of stress included physical illness, fear of infection, and lack of access to resources while in quarantine or isolation settings. Reported indirect sources of stress included social isolation and financial constraints. Levels of trust were generally high regarding mainstream news media, scientific journals, and university-related information sources. For decision-making processes, participants described altered behaviors to socialize safely during the pandemic, which included increased testing, gathering outdoors, and limiting group sizes. Conversely, some undergraduate students reported increases in socialization behaviors after testing negative for COVID-19, while most university employees did not report altered behaviors after negative test results. While some participants described negative feedback regarding university decisions, most feedback for the institution was positive, with participants reporting appreciation for the university’s asymptomatic testing program and other on-campus health response activities.

**Conclusion:** The university’s investment in COVID-related resources, including the asymptomatic testing program and the on-campus quarantine and isolation spaces, were reported to greatly reduce stress and increase perceived safety. Key findings from this research could guide institutional communication, public health protocols, and support for university community members.

## Introduction

The COVID-19 pandemic has resulted in a global public health emergency. The United States reported over 20 million COVID-19 cases and ∼340,000 deaths throughout 2020.^1^ During this time period, Massachusetts recorded 352,558 COVID-19 cases and 12,076 deaths.^2^ This study was conducted at a large public university in Massachusetts during the semester intersession period from December 2020 – January 2021. The epidemic curve among university populations can be viewed in Figure 1.

**Figure 1.**
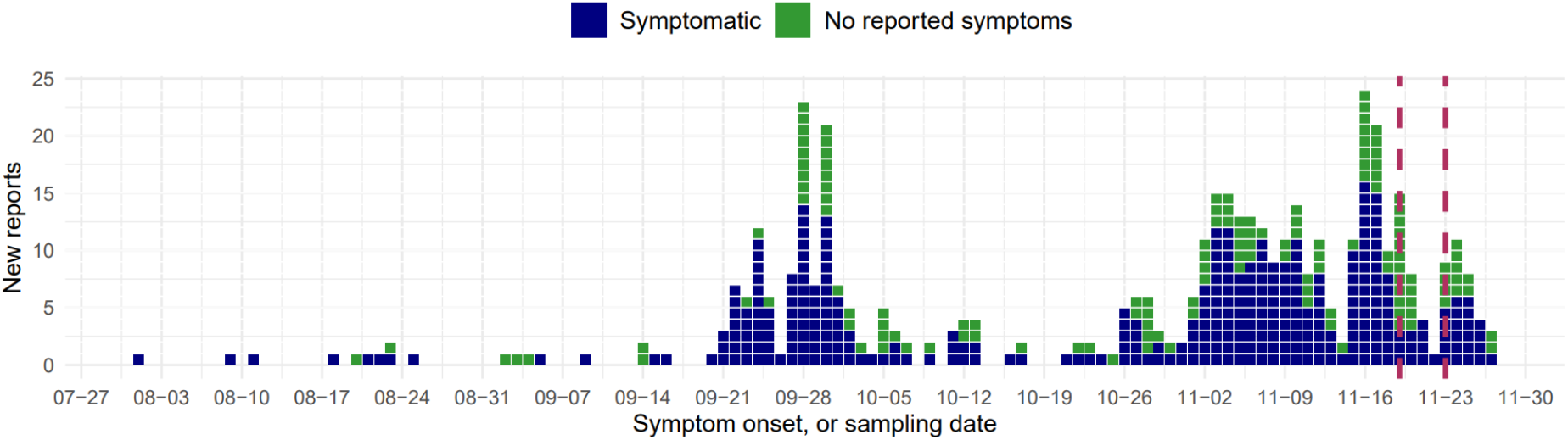
Epidemic curve of COVID-19 diagnoses among university affiliates, August 2020 – November 2020 (Serial intervals show in red).

Few qualitative studies have investigated the experiences of university students during the pandemic. In the UK, mixed-methods designs explored student experiences related to university asymptomatic testing programs, social restriction policies, and quarantine/isolation.^3– 6^ These studies found that asymptomatic testing programs were acceptable to students and increased their perception of safety on campus. Coping strategies for students include peer support, avoiding the news, pursuing hobbies, increased sleep or exercise, and professional mental health care.^3,7–9^ Students requested university-sponsored events to help make connections, especially for newer students.^3–6^ In the UK and US, students expressed a need for clearer institutional communication, with individualized messaging and clear rationale for protocols.^3,4,6,10^

Limited research has examined the COVID-19 experiences of university faculty and staff. Qualitative studies in the UK that included staff perspectives found high satisfaction with testing programs but a significantly increased workload from remote teaching, student accommodations, and supporting student mental health.^3,5^ Employees requested increased student access to health resources, especially for those in quarantine/isolation, and clearer lines of communication for students reporting infections.^3,5^ Staff also reported feeling that on-campus safety was not always prioritized and students under-utilized the resources available to them.^5^ University staff feedback was similar to studies in the general population, with reported concerns about a lack of transparency regarding the evidence used to determine COVID-19 policies.^11^ Finally, the blurring of personal and professional boundaries during the pandemic contributed to lower productivity and missed career opportunities, with disproportionate impacts on women.^12^

Previous studies have had limitations including small sample sizes, limited representation from participants accessing testing programs, over-representation of health-conscious students following policies, and data collection early in the pandemic (July-October 2020).^3–6,12^ The objective of this study was to explore and contextualize student and employee knowledge, attitudes, and practices related to COVID-19 and university COVID-19 policies. These findings may not be applicable to later pandemic waves when perceptions of risk may have changed. This study provides insights relevant to an early stage of the pandemic, which in conjunction with more recent findings may inform future policy related to transmission of SARS-CoV-2 variants or other public health emergencies.

## Methods

### Sample Population and Recruitment

The sample population included adult (≥ 18 years) undergraduate and graduate students, faculty members, and staff members with university affiliation in December 2020. Recruitment occurred via university emails with links to REDCap surveys to assess eligibility, demographics, and consent. Two groups were recruited: focus group discussions (FGDs) consisted of those who did not experience isolation/quarantine during Fall 2020 and interviews consisted of those who experienced isolation/quarantine during Fall 2020. FGD recruitment used a simple random sample of university affiliates obtained from Office of Academic Planning and Assessment. Random samples included 1,200 undergraduates, 100 staff, 100 faculty /librarians, and 100 graduate students. Exclusions for FGD recruitment included administrative positions, anyone on indefinite furlough, and online-only students. For interview recruitment, persons who were contacted by the university Contact Tracing Program in Fall 2020 were eligible. A total of 1,426 FGD recruitment emails and 400 interview recruitment emails were sent, with follow-up reminders at 6-18 days; participants received a $10 merchandise gift card. This study was approved by the University of Massachusetts Amherst IRB (Approval 1873, Nov. 30, 2020).

### Study Context

The Public Health Promotion Center (PHPC) was formed in August 2020 to create and manage the large asymptomatic SARS-CoV-2 testing center across the university, including an in-house contact tracing group and an on-campus quarantine and isolation program. This center was staffed with an interdisciplinary team from across campus, including the College of Nursing, School of Public Health and Health Sciences, Information Technology departments, and the Environmental Health and Safety office. University employees were required to test weekly, while students were required to test twice per week. Students were also required to submit a daily symptom self-check to report any symptomatic illness. Students living either on or off campus were invited to move into on-campus isolation or quarantine residence halls if they tested positive for SARS-CoV-2 or were identified as close contacts, respectively. During the study period several campus scheduling changes occurred, including an earlier start to the semester, minimal holidays,^13^ and “Wellbeing Wednesdays” which consisted of weekly emails with self-care tips.

### Focus Groups and Interviews

FGDs were facilitated by two researchers with separate sessions for undergraduate students and for graduate students, faculty, and staff to avoid power differentials. FGDs including graduate students, staff, and faculty also included a faculty co-facilitator. A total of six FGDs were conducted (Dec 14, 2020 to Jan 15, 2021); each lasted approximately 90 minutes and included 2-5 participants. Four included undergraduate students, while two consisted of graduate students, faculty, and staff. Interviews were conducted individually by one clinically trained faculty researcher. In total fourteen interviews were completed, each lasting 45-60 minutes. FGD and interview guides were semi-structured with use of probes and prompts (Suppplemental Files 1 and 2). Data collection stopped after we exhausted the list of consenting participants and decided that we had a range of attitudes, knowledge and practices.^14^ The number of focus groups and interviews align with sample size recommendations in methodological literature and our use of probing increased the depth of data obtained per person.^15,16^ Only researchers had access to study data (surveys, Zoom links, and transcripts); all files were stored on university-administered secure platforms. The consolidated criteria for reporting qualitative studies (COREQ) guidelines were used throughout this study (Supplemental File 3).^17^

### Data Analysis

Data were analyzed using an inductive thematic approach as in prior work.^4,5,12,18,19^ FGDs and interviews were recorded via Zoom; voice-to-text transcriptions were then cleaned and de-identified by study staff. Analysis was performed using Dedoose software (Version 9.0.17, 2021, Los Angeles, CA). Two researchers (TS, JR) analyzed FGD data to develop a codebook by first reading through transcripts to identify patterns followed by editing organizational style and inductive thematic analysis.^19,20^ Group meetings with the senior research team (SP, SG, AAL) resolved differences and refined the codebook. This codebook was applied to both FGD and interview transcripts. Two researchers independently blind-coded two FGD transcripts and two interview transcripts, with subsequent discussions to ensure consistency in coding. Issues regarding reflexivity were discussed with senior research staff; further details can be found in Supplemental file 4.

## Results

A total of thirteen undergraduate students, one graduate student, three staff members, and five faculty members/librarians participated in the focus groups. Interviews were conducted with nine undergraduates, two graduate students, and three employees. Of these, five participants experienced quarantine, eight experienced isolation, and one experienced both. Demographics are shown in Table 1. Five dominant themes emerged from analysis of these FGDs: COVID-19 knowledge, stress and coping, trust, decision-making, and institutional feedback. These five themes and eighteen sub-themes are shown in Table 2 with representative quotes.

**Table 1.**
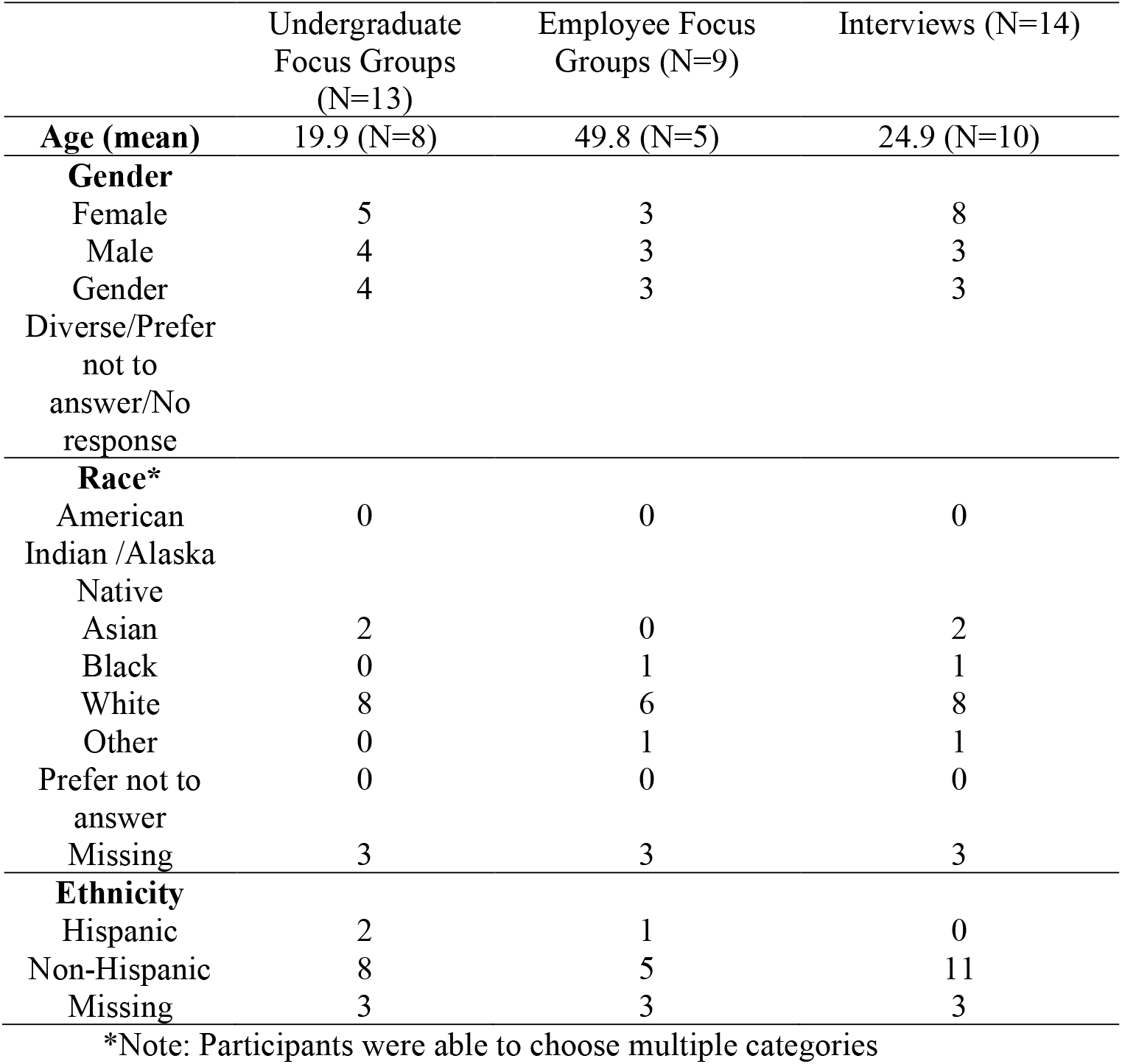
Demographics for focus group and interview participants, COVID-KAP study, 2020 – 2021

**Table 2.**
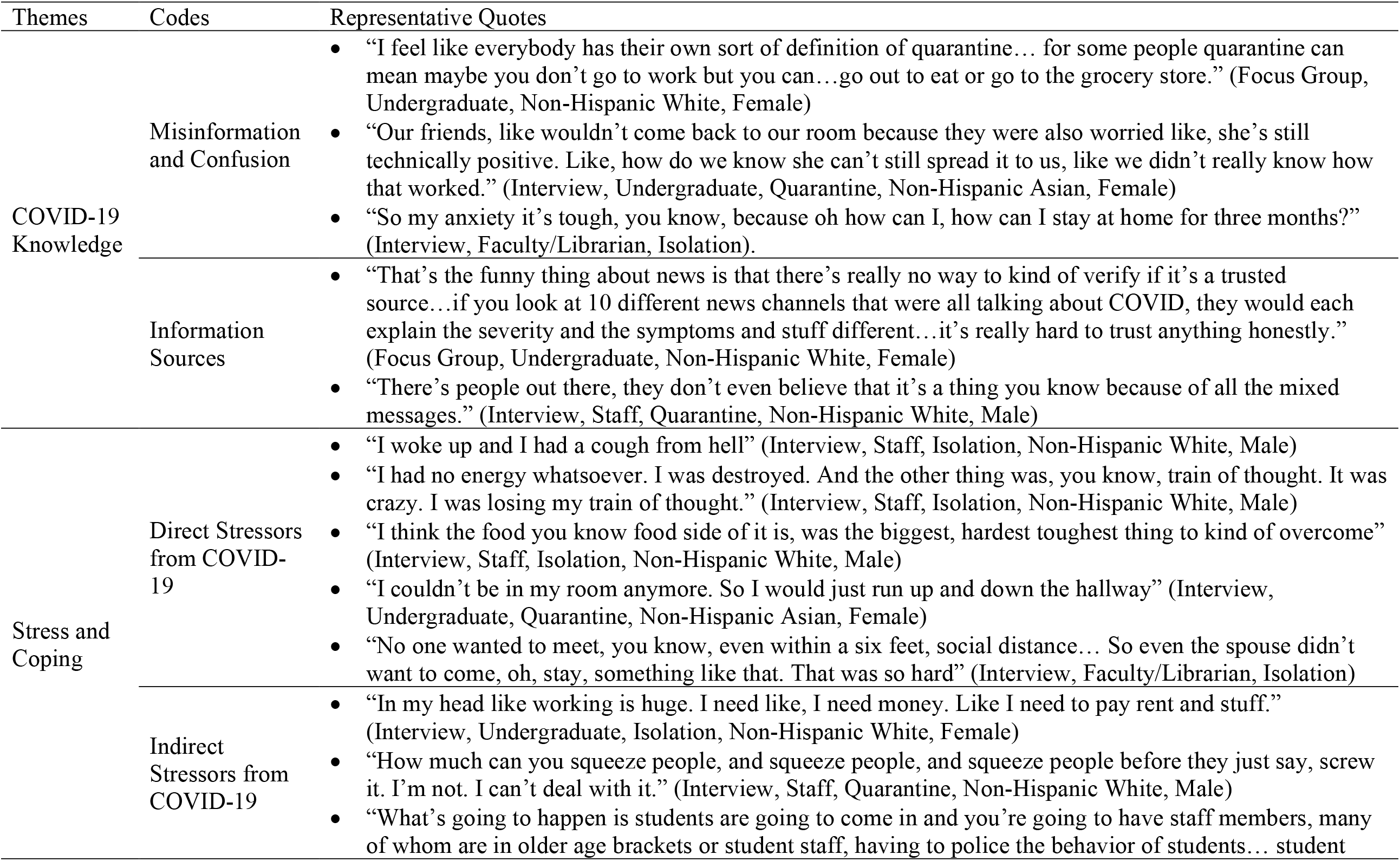
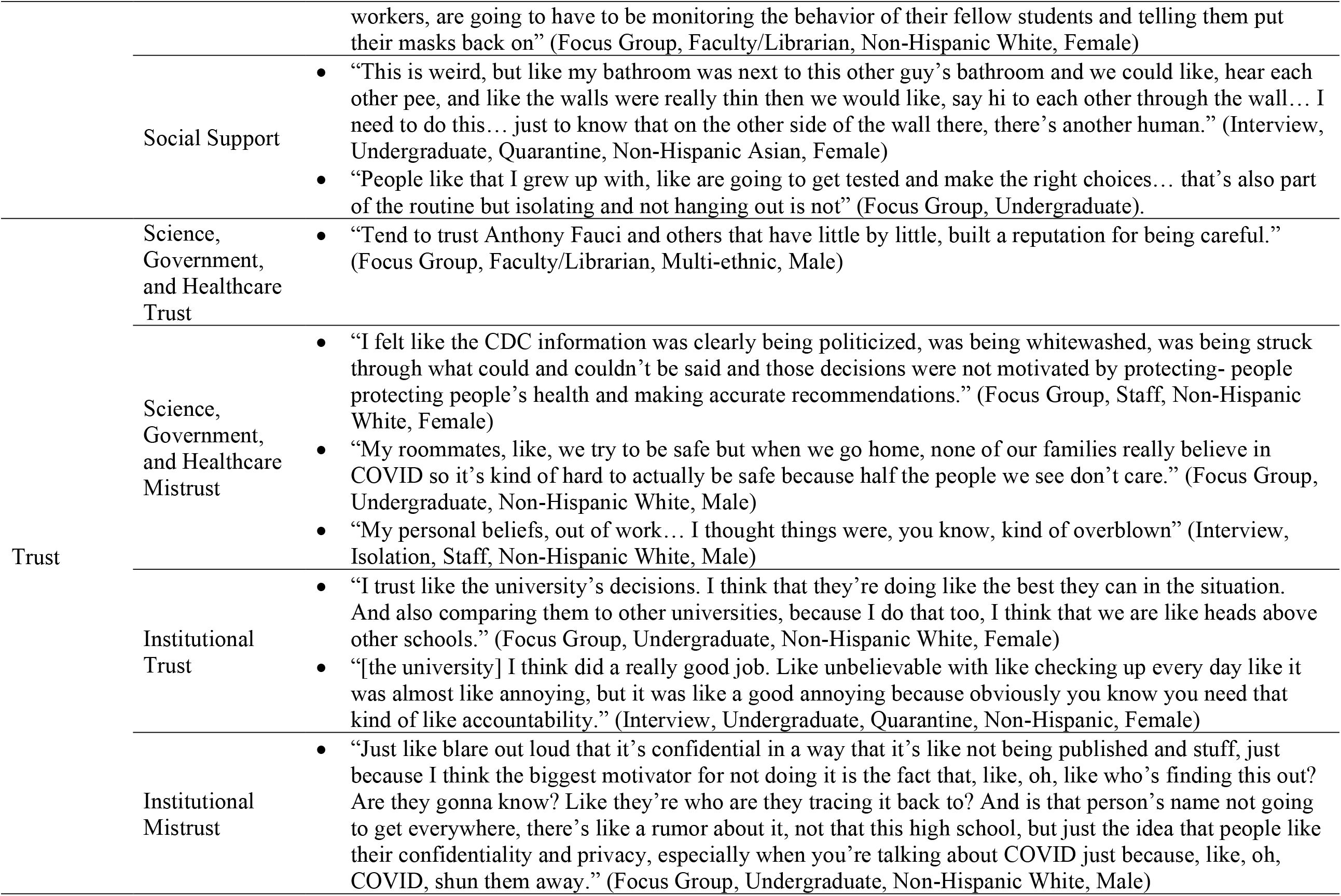
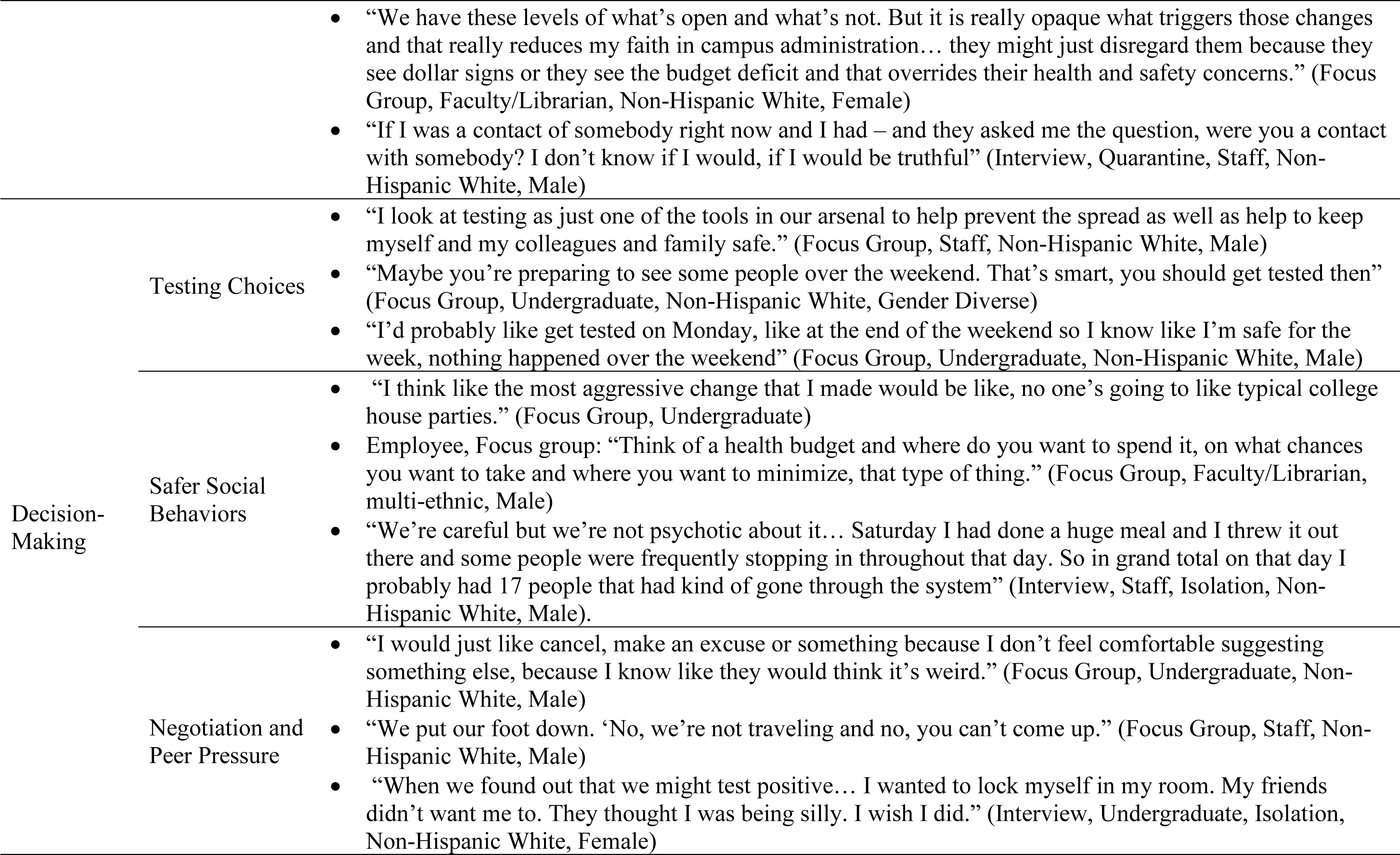
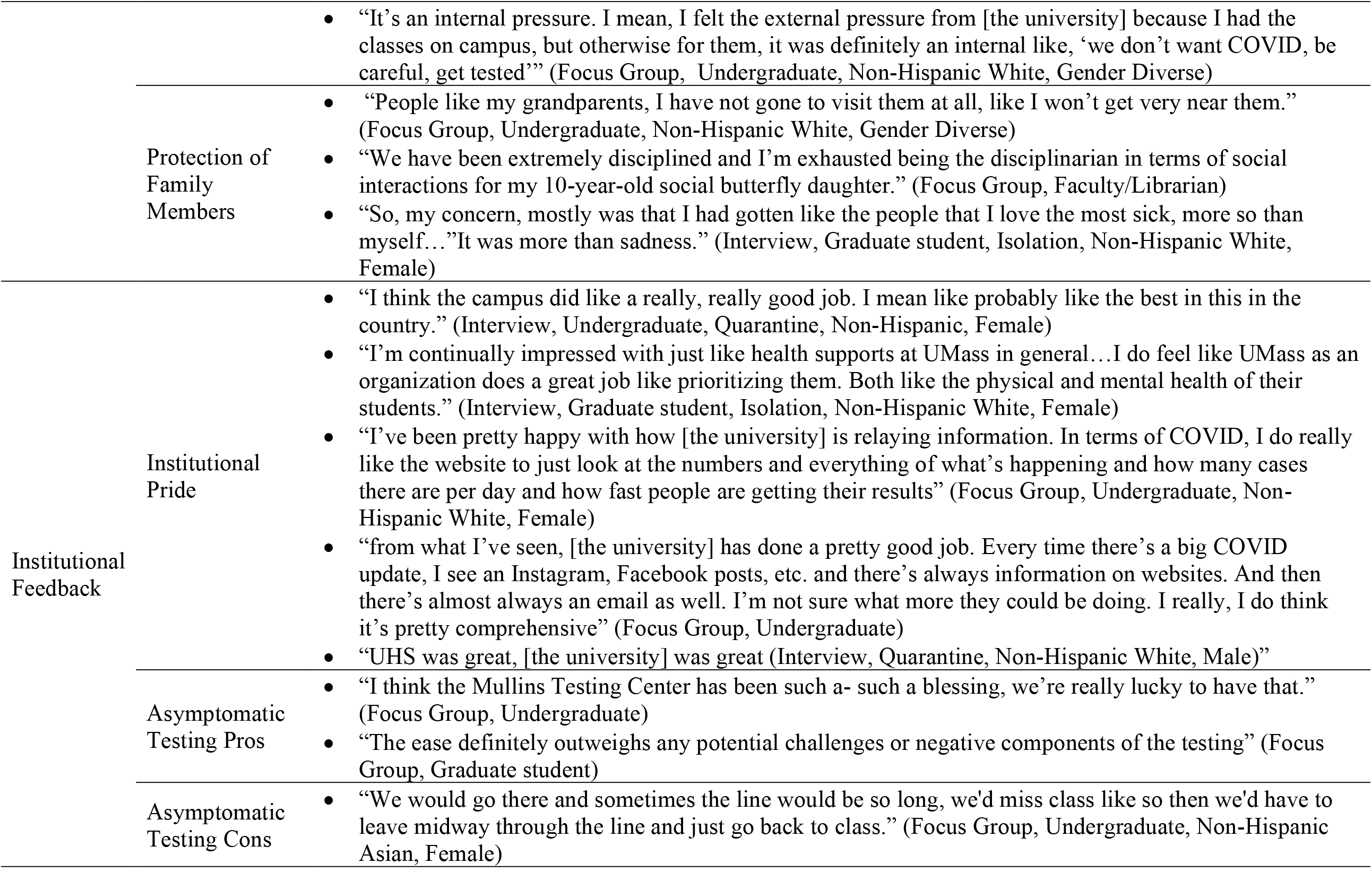
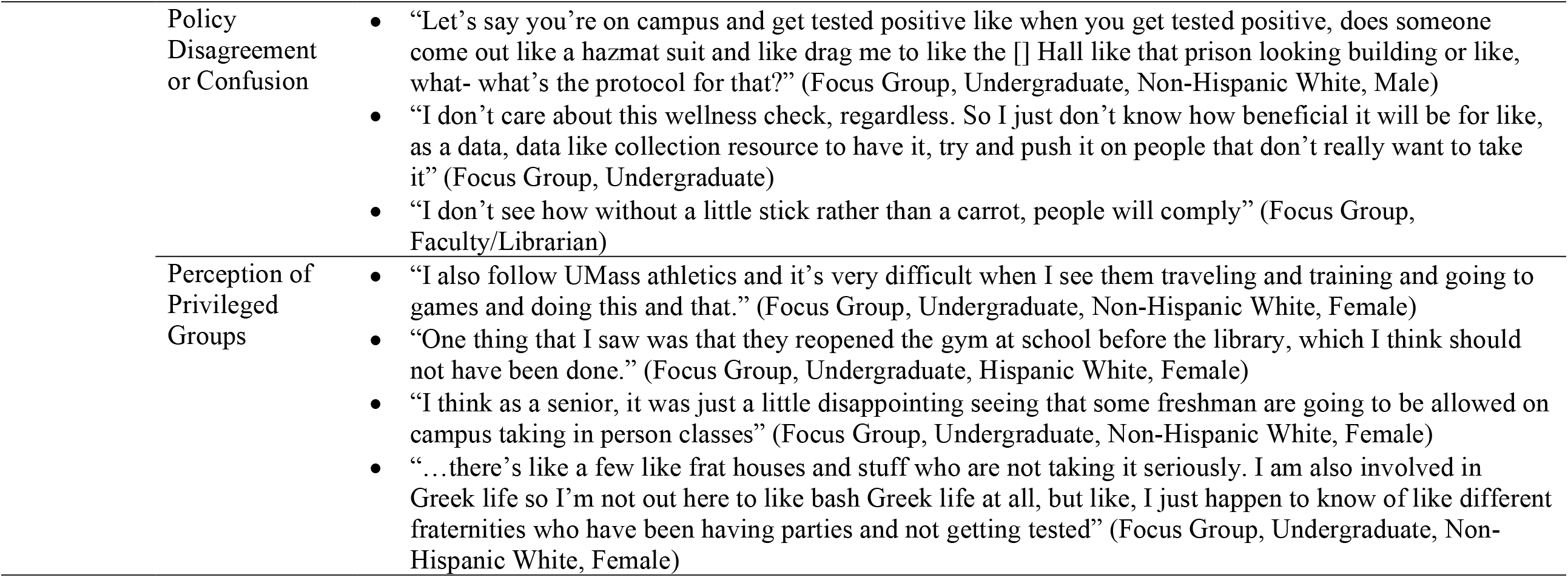
Themes, codes, and their representative quotes for the focus group and interview analyses, COVID-KAP study, 2020 - 2021

### Theme 1: COVID-19 Knowledge

#### Misinformation and Confusion

Focus group participants reported widespread confusion regarding university policies and the underlying science of SARS-CoV-2. Specifically, undergraduates were generally unable to differentiate between ‘isolation’, ‘quarantine’, and ‘social distancing’, and used these terms interchangeably. Most participants indicated widespread COVID-19 misinformation, with students reporting parents as a common source. Moreover, participants were uncertain about delineating “close contact” and believed that everyone in lecture halls or sharing laboratory equipment should be quarantined after a case report. FGD participants also requested more transparency from the university regarding the identities and whereabouts of COVID-positive individuals.

> “I don’t understand why they can’t say where this person was working last and the last time that they were on campus” (FGD, faculty/librarian).

For interviewees, many who experienced isolation believed that their test results were false positives and desired re-testing. Many also expressed confusion about differing time periods for isolation and quarantine; others reported worrying that positive individuals might be infectious after release from isolation. Moreover, the evolving COVID-19 policies were perceived as evidence that there is no right answer.

> “The CDC like went from 14 days to 10 days and they were kind of changing and who knows if we can even trust our government” (Interview, Graduate student, Isolation, Female, Non-Hispanic White).

#### Information Sources

FGD and interview participants commonly cited mainstream news media, government websites, and institutional communications as sources for pandemic information. Several students reported reading original research articles to triangulate information from social media or from friends. In contrast, employees more often discussed seeking information trends within their local communities.

### Theme 2: Stress and Coping

#### Direct Stressors from COVID-19

FGD participants had diverse responses regarding fear of infection, but most reported significant anxiety at the thought of testing positive.

> “I think that people are afraid of getting that like call from like a contact tracer. A fear being like held responsible maybe, for lack of a better term, to like maybe their actions the weekend before” (Focus Group, Undergraduate).

Some undergraduates reported minimal perceived likelihood of severe COVID-19 symptoms, yet many described concern about long-term effects of infection. Employees generally focused on stresses related to family. Interview participants reported poor mental health, particularly in those with pre-existing mental health conditions.

> “I was like in my own bubble, as like a sad person” (Interview, Graduate student, Quarantine, Non-Hispanic Black, Female).

Participants reported decreased productivity but flexible deadlines were reported as protective factors.

> “People were expecting me to give and give and give I’m just like, I can’t right now. I cannot produce at this moment” (Interview, Graduate student, Quarantine, Non-Hispanic Black, Female).

Participants generally reported moderate symptoms, yet certain symptoms lasted for months after the initial diagnosis.

> “My lungs still aren’t great now, I tried to go for a run yesterday, it didn’t go well [4 weeks post diagnosis]” (Interview, Undergraduate, Isolation, Non-Hispanic White, Male).

Confusion regarding COVID-19 policies also exacerbated stress – one student believed they could develop symptoms for 90 days and an employee believed they had to isolate for 90 days. Students staying on campus for isolation/quarantine had no reported concerns regarding food access, but this was reported as a major burden for off-campus students. To mitigate these stressors, participants reported connecting with others electronically, playing video games, watching television, listening to calming music, going for walks, and even talking with strangers through the walls of the on-campus quarantine rooms.

#### Indirect Stressors from COVID-19

Most participants displayed significant stress from indirect consequences of the pandemic which included extensive social isolation, continually disrupted routines, and ongoing financial stress.

> “I’m losing my dang mind… 2020 was the year I learned that I truly am an extrovert, and it is not going well” (Focus Group, Faculty/Librarian, Non-Hispanic White Female).

Participants also reported that the accelerated semester caused additional stress and that workloads for online courses were higher than face-to-face courses. Undergraduates specifically reported that the lack of breaks led to an increase in drinking to cope with stress.

> “I’m gonna have a mental breakdown. So I’m going to just relax my own way, in a sense, and just have people over, get drunk, do other things. So I think, yeah, having a break would have probably minimized how much people get together probably” (Focus Group, Undergraduate, Non-Hispanic White, Male).

Undergraduates mentioned the importance of counseling and suggested expanding access to these resources on campus. Interview participants described the difficulty in meeting deadlines while being sick and many requested extensions for assignments. Employees reported difficulties in ensuring their children remain distanced from others and the need to “police” the behavior of students regarding campus policies. Additionally, several interview participants mentioned guilt for exposing loved ones and stigma associated with being in isolation/quarantine.

#### Social Support

Many, but not all, participants reported altering their socialization behaviors to maintain social connection through increased use of electronic communication and with risk reduction strategies. Other undergraduates reported that testing and small social circles allowed them to socialize with their friends safely. However, other undergraduates emphasized that severely limiting social interactions was not a plausible option.

> “If we get sick, we get sick. None of us really cared” (Focus Group, Undergraduate, Non-Hispanic White, Male).

### Theme 3: Trust

#### Science, Government, and Healthcare Trust and Mistrust

Most participants indicated having strong trust in science, state and federal governments, and the healthcare system.

> “I feel like they’re [CDC] very unbiased, just get straight to the facts” (Focus Group, Undergraduate, Non-Hispanic White, Female)

In contrast, some participants mentioned difficulty in verifying whether COVID-related information is correct, especially from some news sources. Some employees reported feeling like the pandemic was overblown or admitted that they did not abide by all quarantine policies.

#### Institutional Trust

FGD participants generally reported trust in COVID-related information from the university. Some students reported a preference for being called by the contact tracing program instead of informal notifications from friends if they were exposed to COVID-19 to ensure confidentiality and obtain medical advice. Undergraduates reported feeling supported by the university and felt that they received the testing, food, and resources that they needed on campus. Participants who experienced isolation/quarantine described appreciation for staff, food, housing, and counseling resources.

> “I really enjoyed the way that the person [contact tracer] was like, talking to me. Like it felt really genuine and.. it didn’t feel like they were treating me like everyone else” (Interview, Graduate student, Quarantine, Non-Hispanic Black, Female).

#### Institutional Mistrust

Many undergraduates reported that they would prefer to quarantine/isolate without involvement of the Contact Tracing program due to a desire for anonymity, fear of a confidentiality breach, and fear of consequences for failing to adhere to restrictions.

> “I wouldn’t want my name all over like the databases. So that would be like my reason for it, but I know that they just didn’t want to like do all the hassle” (Focus Group, Undergraduate).

Students who experienced isolation reported not always disclosing the names of contacts to contact tracers to avoid making their friends quarantine. Additionally, some FGD participants reported not trusting that the contact tracing process would accurately identify contacts. While most reports of institutional mistrust were directed towards the contact tracing program, other concerns were also mentioned. One student described resistance to technological surveillance that might involve tracking personal movements and was concerned that the university might try to implement these programs. Furthermore, an employee claimed that the university’s COVID-19 dashboard was misleading due to the low positivity rate that results from many asymptomatic tests and described it as “lying with statistics” (Focus Group, Faculty/Librarian, Non-Hispanic White, Female).

### Theme 4: Decision-Making

#### Testing Choices

Participants reported seeking COVID-19 tests for a variety of reasons: to adhere to mandatory testing schedules, after returning from travel, before visiting family, after contact with a positive individual, when pressured by peers, and after perceived high-exposure situations. Multiple students reported that they would not test if it was not free. A concept of ‘proximity testing’ was commonly described by students, where one household member tested to assess the COVID-19 status of the entire household: “One gets tested, we’re good” (Focus Group, Undergraduate). Another student reported that their roommates would assume they were negative if one person in the household tested negative.

> “They decided that it was better that I go get tested for the household and that they’d rather not stand in that line and then maybe get COVID” (Focus Group, Undergraduate, Non-Hispanic White, Gender Diverse).

There was a range of responses to COVID-19 testing results. Some participants reported that negative test results should not result in high-risk behaviors, while others stated that negative test results increased their social behaviors.

> “Definitely makes me want to socialize more. Uh maybe irrational, but when I get a negative test I’ll be like, oh I’m safe now. So I can like, like not be a burden to like older people” (Focus Group, Undergraduate, Non-Hispanic White, Male).

In contrast, employees reported the perception that a negative test should not result in increased socialization.

#### Safer Social Behaviors

Participants described a range of strategies to socialize more safely during the pandemic, which included maintaining a small pod of friends, routine testing, masking and distancing, avoiding crowded places, choosing outdoor locations to socialize, using electronic communication, and delaying visits to high-risk individuals.

> “It’s a balance of like risk tolerance to like so and you don’t really know what your risk tolerance is until like the bad side that you thought you weighed comes true and you’re like, oh shoot, so it was a good it was a good lesson” (Interview, Graduate student, Isolation, Non-Hispanic White, Female)

Perceptions of risk regarding COVID-19 varied among participants; most individuals stated that indoor public areas such as house parties and restaurants were too risky to visit.

Perspectives on what constitutes a “safe” behavior also varied.

> “We were kind of loosey-goosey about it at the beginning of the semester…One of my roommates ended up moving out this semester because she thought we didn’t take COVID seriously” (Interview, Undergraduate, Isolation).

#### Negotiation and Peer Pressure

Many participants described social pressure to engage in activities even when they felt unsafe. However, some reported leaving these situations while others reported lying to avoid unsafe activities.

> “People don’t want to like not seem cool, you know. It’s like if you’re like, cool or whatever, it’s like, oh like, you know, we’ll get together like it’s no big deal. And people just like want to fit in and want to socialize” (Focus Group, Undergraduate, Non-Hispanic White, Female).

Negotiations among social circles were complex, with some students reporting feeling comfortable asking a friend to change plans due to safety concerns while others did not.

> “When it comes to like your friends, it’s a matter of trusting them, I guess, or like trusting that they’re like doing the right thing” (Focus Group, Undergraduate, Hispanic Asian, Female).

Employees reported pressure from family to travel for visits and mentioned having a safe alternative in mind when others suggest unsafe activities. In contrast, peer pressure sometimes led to increased prevention behaviors such as testing.

> “There’s like, major like big group chats that people are a part of… it’s like kind of like aggressively pushed in the big chats anyways and, and online and everywhere. So it’s not really negotiation. It’s like everyone will do it just because they have to” (Focus Group, Undergraduate).

Students in isolation described pressure from friends to not disclose names of close contacts; one student reported peer pressure to avoid quarantine after exposure.

#### Protection of Family Members

Participants described a variety of ways to protect family members including testing before visits, wearing masks, maintaining distance, and meeting outdoors

> “In my house, when my friends come over, they have to wear masks…you have to wear a mask when you’re like walking in the hallway or like walking by my parents. And they’re like ‘what?’ I’m like, ‘yeah, you have to do it’. It’s like those, that’s the only time I guess I’ve convinced my peers, like, do something safe” (Focus Group, Undergraduate, Non-Hispanic White, Male).

Employees discussed protecting their children by using COVID-19 data to determine whether to let them go to school, modifying their own behaviors, and examining children’s social interactions for safety.

### Theme 5: Institutional Feedback

#### Institutional Pride

Participants reported feeling a sense of pride in the university for the asymptomatic testing and contact tracing programs and felt that they provided necessary information and resources throughout the pandemic. Many participants expressed gratitude for free testing and mentioned that other universities required payment for testing.

> “I trust like the university’s decisions. I think that they’re doing like the best they can in the situation. And also comparing them to other universities, because I do that too, I think that we are like heads above other schools” (Focus Group, Undergraduate, Non-Hispanic White, Female).

Participants also reported appreciation for the flexibility to teach and learn remotely.

#### Asymptomatic Testing Center Feedback

Feedback for the university’s asymptomatic testing center was overwhelmingly positive. Participants reported the process was convenient, lines were usually short, results came quickly, and scheduling appointments was simple.

> “I think the whole process is very easy. I don’t really think that there’s necessarily like a hard thing about it. I think it’s like really quick and convenient, you get your results really quick” (Focus Group, Undergraduate, Non-Hispanic White, Female).

Cons for the asymptomatic testing center were overshadowed by the pros. Complaints included issues with the IT portal, occasional long lines, and the unclear layout of the university vs community testing queues.

> “Sometimes the line would be so long, we’d miss class like so then we’d have to leave midway through the line and just go back to class” (Focus Group, Undergraduate, Non-Hispanic Asian, Female).

Participants suggested requiring appointments, more staffing during busy periods, and earlier testing options.

#### Policy Disagreement or Confusion

Participants reported that university policies for testing, wellness checks, consequences for breaking COVID guidelines, and the procedure for being diagnosed with COVID-19 on campus were confusing.

> “It was very unclear. I was going to get tested anyway, but I feel like it was a little bit unclear as to exactly when I need to get tested and was like, like sometimes it said I was out of compliance when I had gotten tested, like the day before. So I think that was a little bit unclear” (Focus Group, Undergraduate, Non-Hispanic White, Gender Diverse).

Other concerns included lack of information on university websites, inability to reach contact tracers, and the lack of breaks throughout the semester. Nearly all participants disliked the Wellbeing Wednesdays and found them unhelpful.

> “Those idiotic ‘Wellbeing Wednesdays’, dear God, those are useless” (Focus Group, Faculty/Librarian).

Students also reported personal hardship from university policies changing abruptly, including restricting access to dining halls and laying off student employees. Additionally, participants reported disliking wellness checks and many refused to fill them out. Furthermore, students and employees described concern about the lack of consequences for those breaking COVID rules.

> “If we want to move those people that are not wearing masks from not wearing them to wearing them, education is not what we’re missing. That we need some figure of authority that tell them ‘sorry, you have to wear a mask’” (Focus Group, Faculty/Librarian, Multi-ethnic, Male)

Staff discussed resentment due to being furloughed and having an increased workload caused by staffing shortages. Additionally, some employees reported frustration with university administrative decisions.

> “I think the biggest problem is that the university administration appears to be making decisions about how to proceed without bothering or without remembering to consult parts of the university. We get these great emails from the Chancellor saying, you know, we’ve talked to stakeholders. That’s nice, I haven’t been talked to. Does that mean I’m not a stakeholder?” (Focus Group, Faculty/Librarian).

Participants who experienced isolation/quarantine reported feeling rushed when having to move out of residence halls, felt interrogated by contact tracers, and thought wellness calls were annoying. One student described the conversation with contact tracers as an interrogation, stating: “’Are you sure you didn’t go to a party’, which I – it was just like of like, I know I didn’t. so like, why are you interrogating me about it?” (Interview, Undergraduate, Isolation).

#### Perception of Privileged Groups

Participants reported the perception that certain groups of individuals received special privileges; on-campus students could access dining halls and computer labs while off-campus students could not. Upperclassmen participants reported that seniors should have in-person courses instead of freshmen. Students described the perception that certain university-sponsored groups were afforded different privileges than others.

> “I just feel like the athletics also, in particular, are held to different standards than the rest of students and I know that everyone loves their sports teams and the athletes and everything but, you know, we should all be following these guidelines and just because you’re an athlete does not mean that you should be getting special privileges that other people aren’t getting” (Focus Group, Undergraduate, Non-Hispanic White, Female).

Furthermore, students and employees reported concern that individuals who did not abide by COVID-19 restrictions were not receiving consequences from the university.

## Discussion

The results of this study highlight the complexities of university affiliates’ experiences during the pandemic that have direct policy implications for other academic institutions. The key findings could guide institutional (1) communication and transparency, (2) implementation of public health protocols, and (3) direct support for university affiliates. This feedback could also foster a more inclusive decision-making process.

A major finding is that institutional communication should be straightforward and consistent, which is essential to building trust and encouraging adherence. While many of study participants reported satisfaction with institutional messaging, we found pervasive misunderstanding of basic terms related to COVID-19 safety protocols. Improved communication regarding guidelines with supporting evidence may encourage compliance with protocols and foster trust in institutional decision-making.^5^ Indeed, some participants interpreted the lack of clarity as a sign of arbitrary rules and loose safety protocols that were easily adaptable to individual preferences. While some of these changes were due to the rapidly evolving understanding of the virus, it led to some mistrust. At its worst, misinformation contributed to disbelief in positive SARS-CoV-2 test results or increased stress, as was found in other studies.^4– 6,11^ Lack of transparency about repercussions led to dishonesty with contract tracers about close contacts and generated frustration in both students and employees who were following public health guidance. Institutional communications could be improved by beginning with empathy, emphasizing the goal of safety, and keeping messages as simple as possible.^4,10^ Institutions may also need to tailor messaging to various roles on campus and acknowledge the added vulnerability of essential staff and low-income or international students.^4,10^

The rapid implementation of public health protocols during the fall 2020 semester was often unable to adequately balance the immediate public health response (reducing COVID-19 transmission) with broader health goals (ensuring the well-being of everyone impacted). While the altered semester schedule aimed to reduce transmission from travel, the consequent lack of breaks contributed to increased workloads and reported mental health concerns. Participants did not find “Wellbeing Wednesdays” useful for reducing stress. Moreover, a lack of information about the evidence informing decisions contributed to participants’ mistrust in the university, similar to findings from a large population-based survey of public perceptions.^11^

Investment in direct support for university affiliates contributed to reported student and employee resilience. Many interview participants expressed gratitude for university support during quarantine/isolation. Similar to feedback in other studies, students who resided on-campus for quarantine/isolation appreciated the free accommodations and meal deliveries.^5^ The asymptomatic testing program reportedly fostered a perception of safety on campus and reduced individuals’ COVID anxiety, similar to another study.^3^ Many undergraduates reported gratitude towards professors for accommodations during a stressful semester; together, these responses reinforce the widespread effects of institutional investment in programs to support university affiliates.

However, a lack of institutional support may exacerbate stress and mental health challenges among community members.^8,13^ Instructors reported the combination of increased family duties and students with additional needs contributed to an unsustainable workload. Similar concerns have been raised in other studies, with employees requesting a continuation of flexible pandemic policies and using the momentum of change to improve other inequitable policies.^3,5,12^ University participants have reported increased physical exhaustion and anxiety or fear about both the personal health and interpersonal relationship consequences of a positive test.^3,4,21^ One study found that university students who tested positive for COVID-19 were more likely to experience food insecurity or mental health disorders such as anxiety and depression.^22^ Some of the coping strategies mentioned included spending time outdoors, exercising, socializing, and distractions, findings similar to other studies.^3,5,21^ The importance of social support was highlighted in the decision-making theme of our results and has come up many times in other studies.^3–5,12^ Institutions might leverage the importance of social support to encourage community members to support each other during times of crisis.

Findings from this study suggest a need for broader institutional policies that avoid perceptions that certain groups have greater privileges or that enforcement is inconsistent. Students reported fewer incentives for off-campus students to participate in testing because there was no reward for doing so (i.e. they still could not use the gym or dining hall). Some students also reported that certain university-sponsored groups appeared to have special privileges during the pandemic and that the gym was re-opened before the library. Together, these were construed as the institution valuing athletics over academics. Staff members felt that their jobs and safety were not prioritized due to the furloughs and expectation that most staff work on-campus throughout the pandemic, which was also found in another study.^5^ Other studies have found that mental health challenges and stress are higher in females, economically disadvantaged students, and people with personal experiences with COVID-19.^3,4,7^ In summary, if policies are not applied and enforced equitably, they may contribute to reduced well-being.

### Strengths and Limitations

This study’s strengths included a rigorously designed study, timely data collection, and inclusion of diverse perspectives. The study was conducted at a large university which experienced several COVID-19 case surges during the fall 2020 semester, making results more generalizable than studies conducted in lower transmission settings.^3^ Our participants had broad experience with COVID-19 protocols and were therefore able to provide feedback about how to improve processes. Additionally, interviews for those who experienced isolation/quarantine were conducted by a researcher with no involvement in COVID response on campus, increasing the validity of our data because participants likely felt more comfortable giving honest feedback. We also gathered feedback from participants across a spectrum of caution and compliance with COVID-19 protocols, addressing a limitation of previous studies which mainly included compliant participants.^5,6^

One limitation of this study was a modest response rate to recruitment emails. Though we sent reminder emails, our response rate was 1.5% and 3.5% for focus groups and interviews, respectively. This may have been due to the timing of recruitment, which was during the intersession period. A few of our focus groups only included 2-3 participants, which may have reduced the benefits of group dynamics.

## Conclusions

The results of this study describe the profound amount of direct and indirect stress that university affiliates have faced during the pandemic. Investment in university resources, such as the asymptomatic testing program and the on-campus quarantine and isolation spaces, can greatly decrease stress and increase perceived safety. Additionally, the limited levels of knowledge regarding COVID-19 presents an educational opportunity for university populations. Overall, the key findings from this research could guide institutional communication and transparency, public health protocols, and support for university community members.

## Data Availability

The datasets generated and analyzed during the current study are not publicly available in order to preserve participant confidentiality. Data may be available from the corresponding author upon reasonable request.

## Acknowledgements

We would like to express our gratitude to everyone who contributed to this study. Research assistants Eva Chow and Meghan Fernandes completed the transcription and clean-up for interviews. The Public Health Promotion Center Directors Ann Becker and Jeffrey Hescock provided funding and support.

## Declarations

### Ethics approval and consent to participate

This study was approved by the University of Massachusetts Amherst Institutional Review Board (Approval 1873, Nov. 30, 2020). Informed consent was obtained electronically from all participants.

### Authors’ Contributions

Conceived the study questions and design (SP, SG, AL, TS, JR). Collected data (JR, TS, KW, SG, AL, SP). Analyzed the data (JR, TS, SG, AL, SP). Drafted the manuscript (TS, JR). Reviewed and approved the final version (JR, TS, KW, SG, AL, SP).

## Supplemental Materials

Supplemental file1 (Word Document .docx) - Focus Group Guide Supplemental file 2 (Word Document .docx) – Interview Guide

Supplemental file 3 (Word Document .docx) - Consolidated criteria for reporting qualitative studies (COREQ): 32-item checklist

Supplemental file 4 (Word Document .docx) - Description of codebook themes and codes

## References

1. Cumulative Cases. Johns Hopkins Coronavirus Resource Center. Accessed March 30, 2022. https://coronavirus.jhu.edu/data/cumulative-cases

2. Massachusetts Department of Public Health COVID-19 Dashboard. Weekly COVID-19 Public Health Report, Thursday December 31, 2020. https://www.mass.gov/doc/weekly-covid-19-public-health-report-december-31-2020/download

3. Blake H, Corner J, Cirelli C, et al. Perceptions and Experiences of the University of Nottingham Pilot SARS-CoV-2 Asymptomatic Testing Service: A Mixed-Methods Study. Int J Environ Res Public Health. 2020;18(1):188. doi:10.3390/ijerph18010188

4. Blake H, Knight H, Jia R, et al. Students’ Views towards Sars-Cov-2 Mass Asymptomatic Testing, Social Distancing and Self-Isolation in a University Setting during the COVID-19 Pandemic: A Qualitative Study. Int J Environ Res Public Health. 2021;18(8):4182. doi:10.3390/ijerph18084182

5. Knight H, Carlisle S, O’Connor M, et al. Impacts of the COVID-19 Pandemic and Self-Isolation on Students and Staff in Higher Education: A Qualitative Study. Int J Environ Res Public Health. 2021;18(20):10675. doi:10.3390/ijerph182010675

6. Wanat M, Logan M, Hirst J, et al. Perceptions on Undertaking Regular Asymptomatic Self-Testing for COVID-19 Using Lateral Flow Tests: A Qualitative Study of University Students and Staff. Public and Global Health; 2021. doi:10.1101/2021.03.26.21254337

7. Moriarty T, Bourbeau K, Fontana F, McNamara S, Pereira da Silva M. The Relationship between Psychological Stress and Healthy Lifestyle Behaviors during COVID-19 among Students in a US Midwest University. Int J Environ Res Public Health. 2021;18(9):4752. doi:10.3390/ijerph18094752

8. Kaur J, Chow E, Ravenhurst J, et al. Considerations for Meeting Students’ Mental Health Needs at a U.S. University During the COVID-19 Pandemic: A Qualitative Study. Front Public Health. 2022;10. Accessed March 30, 2022. https://www.frontiersin.org/article/10.3389/fpubh.2022.815031

9. Wallace KF, Putnam NI, Chow E, Fernandes M, Clary KM, Goff SL. College students’ experiences early in the COVID-19 pandemic: Applications for ongoing support. J Am Coll Health. 2021;0(0):1–10. doi:10.1080/07448481.2021.1954011

10. Mackert M, Table B, Yang J, et al. Applying Best Practices from Health Communication to Support a University’s Response to COVID-19. Health Commun. 2020;35(14):1750–1753. doi:10.1080/10410236.2020.1839204

11. Enria L, Waterlow N, Rogers NT, et al. Trust and transparency in times of crisis: Results from an online survey during the first wave (April 2020) of the COVID-19 epidemic in the UK. Duse AG, ed. PLOS ONE. 2021;16(2):e0239247. doi:10.1371/journal.pone.0239247

12. Gottenborg E, Yu A, Naderi R, et al. COVID-19’s impact on faculty and staff at a School of Medicine in the US: what is the blueprint for the future? BMC Health Serv Res. 2021;21(1):395. doi:10.1186/s12913-021-06411-6

13. Revised Academic Calendar for Fall 2020 | UMass Amherst Spring 2022. UMass Amherst Spring 2022 : UMass Amherst. Accessed February 18, 2022. https://www.umass.edu/coronavirus/news/revised-academic-calendar-fall-2020

14. Nelson J. Using conceptual depth criteria: addressing the challenge of reaching saturation in qualitative research. Qual Res. 2017;17(5):554–570. doi:10.1177/1468794116679873

15. Weller SC, Vickers B, Bernard HR, et al. Open-ended interview questions and saturation. Soundy A, ed. PLOS ONE. 2018;13(6):e0198606. doi:10.1371/journal.pone.0198606

16. Nascimento L de CN, Souza TV de, Oliveira IC dos S, Moraes JRMM de, Aguiar RCB de, Silva LF da. Theoretical saturation in qualitative research: an experience report in interview with schoolchildren. Rev Bras Enferm. 2018;71(1):228–233. doi:10.1590/0034-7167-2016-0616

17. Tong A, Sainsbury P, Craig J. Consolidated criteria for reporting qualitative research (COREQ): a 32-item checklist for interviews and focus groups. Int J Qual Health Care. 2007;19(6):349–357. doi:10.1093/intqhc/mzm042

18. Mowbray F, Woodland L, Smith LE, Amlôt R, Rubin GJ. Is My Cough a Cold or Covid? A Qualitative Study of COVID-19 Symptom Recognition and Attitudes Toward Testing in the UK. Front Public Health. 2021;9:716421. doi:10.3389/fpubh.2021.716421

19. Braun V, Clarke V. Using thematic analysis in psychology. Qual Res Psychol. 2006;3(2):77–101. doi:10.1191/1478088706qp063oa

20. Crabtree B, Miller W. Doing Qualitative Research (Research Methods for Primary Care). 2nd ed. SAGE Publications, Inc.; 1999.

21. Frontiers | Considerations for Meeting Students’ Mental Health Needs at a U.S. University During the COVID-19 Pandemic: A Qualitative Study | Public Health. Accessed February 18, 2022. https://www.frontiersin.org/articles/10.3389/fpubh.2022.815031/full

22. Self-reported COVID-19 infection and implications for mental health and food insecurity among American college students | PNAS. Accessed February 18, 2022. https://www.pnas.org/content/119/7/e2111787119

