## Supplemental File 1 for "Knowledge, attitudes, and practices regarding COVID-19 among university students and employees in Massachusetts, USA: a qualitative study"

**Supplemental file 1**: Focus Group Guide

**Focus Group Discussion Guide**

Notes for researchers:

- Throughout discussion, make a list of all questions or mis-information that is heard and address it at the end of the session.
- Do not say “great” to someone’s response - say **“thank you very much for sharing that” +/- “does anyone else have thoughts about this”**? Or using the probes to keep conversation flowing while using neutral words and tones.
- Post simple version of questions in the Chat in case people forget
- If everyone appears to agree with one person’s view, ask, “**Are there any other thoughts? Does anyone think differently about this?”**
- it is good practice after each question block to say, **“Before we move on to the next question, I just want to take a minute to summarize my understanding of your responses. Please let me know if I missed anything or if my understanding is off.**" This gives you a pause to collect your thoughts, let you and the notetaker be sure you covered all you wanted to and/or followed up on responses where you felt there might be more if you probed.

[Secondary Moderator] Hi everyone and thank you for agreeing to talk with us about your experiences this semester related to the pandemic and UMASS’s response.

My name is [ fill in name of researcher] and I am a [primary role and department at university]. The other co-lead for this focus group is [fill in name of researcher] and [pronouns] is a [primary role and department at university].

We are working with a team that is trying to learn how UMASS can do the best job possible supporting the UMass community and keeping everyone on campus and off safe during this really challenging time.

This focus group will aim to let everyone have a chance to share their thoughts together about the campus Public Health Response and gather information to help inform Spring 2021 planning for the asymptomatic testing program, campus education campaigns, and the isolation and quarantine resources provided.

[show Ground Rules slide] – Secondary Moderator Pull Up

[Lead Moderator] Hi everyone. I would like to start by going over some Ground Rules for the focus group discussion today.

- First, only one person should speak at a time, so please keep yourself on “Mute” when you are not speaking. The “Chat” function at the bottom of your Zoom screen can be used to post questions or comments when others are speaking.
- Next, please be respectful of one another’s opinions and allow everyone to get a chance to speak.
- Finally, all information shared here will be anonymized and will be treated as confidential. All participants are asked to hold what is said in this room as confidential and to not share the details. Please do not take any photos or screenshots during the session.
- If at any point you do not want to continue participating in this discussion, you are free to ask to stop, skip a question, or leave the zoom session and we will not contact you further regarding this study.
- The purpose of this information-gathering is to understand the UMass community’s  experience during the pandemic and to help inform the decisions of the Public Health Promotion Center team for Spring planning.
- Just a reminder, we will be recording this session, with the recordings accessible only to research staff. If you feel uncomfortable with this you may leave at any time. Are there any questions regarding the use of zoom or our plan for this session?

[answer any questions – start recording at this time]

[Lead Moderator] Let’s begin by sharing a little bit about ourselves. I would like to go around the room and have everyone share their name, role at UMass/department, and your favorite pandemic activity. Let me start as an example: My name is [fill in name], I am a [role at UMass/department] and my favorite pandemic activity is [fill in activity].

[put question in the Chat and announce that question can be viewed in the Chat]

- In chat: Name, role at UMass/department, favorite pandemic activity

[go around the “room” and call on people one at a time]

[Lead Moderator] We are interested to know where the campus community hears about health information and what sources you find easiest to access and the most reliable or trusted.

In chat: What sources do you use for health information - which do you find easiest to access/most reliable or trusted.

- Could you tell us a little about where you usually get information about COVID?
  - **Probe**: why there? What makes you trust that source?
  - Where do you think your friends/colleagues get their information?
    - If you are asked a question about COVID, what do you do?
    - If your friend had or shared incorrect information about COVID, would you feel comfortable talking with them about it?
  - Have you relied on any UMass resources to learn about COVID? If so, which ones? Which resources helped you most? Is there anything more you would like to see?
  - Do you follow the Umass IG or Twitter accounts? Which ones? What do you think about them? Why?

[Follow-up questions]

- The process of contact tracing can be confusing. What have you heard about contact tracing?
  - What else have you heard? What do your friends think about it?
- How do you feel about health staff potentially calling you or your friends/family/contacts?
- Could you tell me what you know or have heard about quarantine?
  - **Probe**: where did you hear that? Why do you think that’s the case? Can you tell me more? What do you hear from your friends?
- Could you tell me what you know or have heard about isolation?
  - **Probe**: where did you hear that? Why do you think that’s the case? Can you tell me more? What do you hear from your friends?
- What do you think it would be like for you if you tested positive?

[Lead Moderator] We are interested to know more about your experiences regarding asymptomatic testing at the Mullins Center. What is one easy thing and one hard thing about getting an asymptomatic COVID-19 test at Mullins?

In chat: One easy thing & one hard thing about getting an asymptomatic COVID-19 at Mullins?

[Follow-up questions]

- Could you suggest any improvements that could be made at the Mullins Center that would make testing easier?
- How do you decide when to get a test?
  - If you think you were possibly exposed, what would you do?
- Does getting a negative test change any of your decisions about how you socialize?

Other probes if appropriate: [mostly for Undergrads]

- How do you define health? - [might be helpful probe if someone doesn’t think COVID is something people their age need to worry about]

[Lead Moderator] Socializing is so important to all of us as human beings. Can you share about some of the ways you’ve been socializing or meeting your needs to socialize? It is OK to share even if this isn’t within the current guidelines or recommendations. We will keep what is said here confidential. No shame or blame - we are just seeking some understanding.

In chat: The ways you’ve been socializing or meeting your needs to socialize?

[Follow-up questions]

- How do you decide which activities might be safe or less safe?
- Do you use any strategies to minimize risk? **Probe**: can you tell me more?
- Are there some activities that you choose to avoid because they are too risky?
- Do you feel comfortable asking a friend to do something different if you are uncomfortable with potential COVID risk?
- Do you and your friends or social groups discuss COVID risk, what are those discussions like?
- If roommates come up - What would help you and roommates feel prepared and protected against COVID?

Potential probes to add if parties come up: [for Undergrads]

- If you have had parties or in general, do you register your parties or believe that registering parties would be effective?
- Have out of state students or students from other college campuses come to parties? If so, how did that make you feel?
- If you are unhappy with parties occurring, what could UMass do to help?

[Lead Moderator] Thank you all for sharing about your individual experiences. Let’s turn the discussion now to thinking about the broader campus view. What do you feel has been going well in the decisions the campus has made regarding COVID-related planning?

In chat: What has been going well in the decisions the campus has made regarding COVID-related planning?

[Follow-up questions]

- Are there specific things that you think were not great decisions or that you see are not working?
- Is there anything else we haven’t covered that you’d like to share about any part of your experience this Fall related to the Public Health Promotion Center?

[Secondary Moderator] Thank you all for your participation in this focus group discussion. This is the end of the discussion topics that we wanted to cover. We will now talk a little bit about questions that came up during the discussion.

[go through list of questions or misinformation and try to clarify]
