## Supplemental File 2 for "Knowledge, attitudes, and practices regarding COVID-19 among university students and employees in Massachusetts, USA: a qualitative study"

**Supplemental file 2**: Interview Guide

**Interview Guide:**

Hello and thank you for agreeing to talk with us about your experiences this semester related to the pandemic and UMASS’s response. My name is [Name] and I am a [*PH/nursing/other student]* working with a team that is trying to learn how UMASS can do the best job possible supporting students and keeping everyone on campus and off safe during this really challenging time.

Information shared here will be anonymized and will be treated as confidential. If at any point you do not want to continue participating in this discussion, you are free to ask to stop, skip a question, or leave the zoom and we will not contact you further regarding this study. The purpose of this information-gathering is to understand the student experience in Isolation and Quarantine and to help inform the decisions of the Public Health Promotion Center team for Spring planning and to share actionable feedback items with campus administration as they arise. The goal is to learn from your day to day experiences as a person who was *isolated/ quarantined* and to capture information about your experience. I want you to know, and I will remind you that nothing you say here will get you or anyone else in trouble or be used for any disciplinary purpose so be as honest and open as you’d like.

1) I’d like to start by asking if you can tell me a little bit about you. [Probes: undergrad/grad; year; major if one; living on or off/ housing situation; local support systems, general demographics]
   a. What did you know about the contact tracing process before this experience?
   b. How did you feel about it before you experienced it yourself?
2) Thank you. Next, I’d like to give you an opportunity to share about your experience overall in *Isolation OR Quarantine.*

Potential Probes/ Follow-ups:

- To begin, tell me a little about how you felt when you received the initial call with the news that you needed *Isolation or Quarantine.*
  - What worked well in this first call?
  - What would you recommend changing about this process (if anything)?
  - What did you understand about your instructions to be based on this first call?
  - Did you feel you had enough information? Why or why not?
- Next, I’d like you to share about the days you spent in *Isolation/ Quarantine.*
  - Were there specific parts of the experience that were positive/helpful? (daily calls, testing visits, food delivery)
  - Were there things that happened that weren’t ideal or you think could be improved?
  - The pandemic in general has been stressful for many people and being In isolation/quarantine can be even harder. Can you tell me a little bit about how the experience may have impacted your mental health?
  - The University has identified resources, such as referral to the Center for Psychological Health to help people cope with the experience of Isolation and Quarantine.
    - What resources did you hear about while in *Isolation or Quarantine*?
    - Did you use any of the resources?
      - Can you tell me about your experience with the resource? (positives/ negatives/ suggestions for improvement)
      - What else would have helped you?
  - How was your academic experience affected by *Isolation or Quarantine?*
    - Can you tell me about any supports you received? What about supports you would have liked to have?
    - What did you find most difficult about managing the academic aspects of Q&I?
  - *Isolation / Cases ONLY :* The coronavirus has made some people quite sick physically. Could you tell me a little about your physical experience with the virus?
    - What supports, follow up, or care did you receive for your physical health?
    - Do you have comments or suggestions on how better to support folks who are sick or experiencing symptoms while in isolation?
    - What did you find most helpful during any difficult time periods?
  - Knowing that this interview will be held confidentially, I wanted to ask about your experience following the instructions and recommendations from the Public Health team.
    - What parts of the instructions were easy for you to always follow?
    - What parts were more difficult?
    - Did you have to make any hard decisions? Can you tell me more about it?

3) Next, I’d like to hear about how well you feel UMASS has been handling the COVID situation generally. [What do you feel has been working well/good decisions; what do you see as opportunities for improvement in spring semester?]

- What do you know about the pandemic response at the University?
  - How did you learn what you know?
  - How do you feel about the University responses?
- Do you think your friends know where to find information? Where do they look for information?
- What do you know about the University guidance for gatherings? What do you think your peers or students are doing related to this guidance?
  - How do you feel about this guidance, can you tell me more?
- What do you know or what have you heard about the asymptomatic testing program?
  - How do you feel about getting regular testing?
- Have you or your friends had questions about testing or public health issues? Are there things related to the pandemic that you’d like to learn more about?

4) Is there anything else we haven’t covered that you’d like to share about any part of your COVID experience?
