## Supplemental File 3 for "Knowledge, attitudes, and practices regarding COVID-19 among university students and employees in Massachusetts, USA: a qualitative study"

**Supplemental file 3:** Consolidated criteria for reporting qualitative studies (COREQ): 32-item checklist

| Topic and Item | Guide Questions | Response |
| --- | --- | --- |
| **Domain 1: Research team and reflexivity** | | |
| *Personal Characteristics* | | |
| Interviewer/facilitator | Which authors conducted the interviews and focus groups? | Interviews – Sheila Pennell (SP)  Focus Groups – Johanna Ravenhurst (JR), Teah Snyder (TS), Kate Wallace (KW), Andrew Lover (AL), Sarah Goff (SG) |
| Credentials | What were the researcher’s credentials? | SP: RN & PhD in Nursing  JR: MSPH in Tropical Medicine  TS: MPH in Epidemiology and Biostatistics  KW: BS + MPH in Public Health expected May 2022  AL: MPH, PhD in Epidemiology  SG: MD, PhD in Public Health |
| Occupation | What was their occupation at the time of the study? | SP: Clinical Assistant Professor  JR: MS student in Epidemiology  TS: PhD candidate in Epidemiology  KW: MPH student in Epidemiology  AL: Assistant Professor in Epidemiology  SG: Associate Professor, Health Policy and Management |
| Gender | Was the researcher male or female? | 5 females, 1 male |
| Experience and training | What experience or training did the researcher have? | SP: clinically trained nurse, experienced interviewer  JR, TS, KW: Mixed-methods research students, trained interviewers  AAL: Mixed-methods researcher, experienced interviewer.  SG: clinically trained physician, experienced interviewer, mixed-methods researcher |
| *Relationship with participants* | | |
| Relationship established | Was a relationship established prior to study commencement? | Researchers met participants during focus groups or interviews. SP knew one employee prior to the interview (colleague) |
| Participant knowledge of the interviewer/facilitator | What did the participants know about the researchers? (e.g. personal goals, reasons for doing the research) | Participants were aware that researchers were a team from the School of Nursing, the School of Public Health and Health Sciences, and the Public Health Promotion Center. They knew that researchers were affiliated with the university, and they had our names and position titles. |
| Interviewer characteristics | What characteristics were reported about the interviewer/facilitator? (e.g. bias, assumptions, reasons and interests in the research topic) | Participants knew researchers had been working on COVID-related efforts and wanted to know what was working well and where we could do better |
| **Domain 2: Study design** | | |
| *Theoretical framework* | | |
| Methodological orientation and theory | What methodological origination was stated to underpin the study? (e.g. grounded theory, discourse analysis, ethnography, phenomenology, content analysis) | Thematic analysis |
| *Participant selection* | | |
| Sampling | How were participants selected? (e.g. purposive, convenience, consecutive, snowball) | Interviews – purposive sampling of people who had experienced isolation/quarantine at institution  Focus groups – purposive and random sampling – used a random sample of people from roles of undergraduate, graduate students, and faculty/staff in order to gain insights from a variety of perspectives |
| Method of approach | How were participants approached? | email |
| Sample size | How many participants were in the study? | Interviews – 14 participants  Focus groups – 22 participants |
| Non-participation | How many people refused to participate or dropped out? Reasons? | Interviews – 15 scheduled but did not show up for interview.  Focus groups – 15 scheduled but did not show up for focus group. Many emailed in advance to say they couldn’t make it, some were no-shows. |
| *Setting* | | |
| Setting of data collection | Where was the data collected? | Data were collected online (Zoom)  Interviews were 1-on-1 Focus groups were divided based on role (undergraduate; faculty/staff/graduate student) |
| Presence of non-participants | Was anyone else present besides the participants and researchers? | No |
| Description of sample | What are the important characteristics of the sample? (e.g. demographic data, date) | Interviews: Age, gender, race, ethnicity, quarantine/isolation, role at institution  Focus groups: Age, gender, race, ethnicity, role at institution |
| *Data collection* | | |
| Interview and focus group guide | Were questions, prompts, guides provided by the authors? Was it pilot tested? | Yes – both interview and focus group guides are provided in supplemental materials No – the guides were not pilot tested |
| Repeat interviews | Were repeat interviews carried out? If yes, how many? | No repeat interviews or focus groups. |
| Audio/visual recording | Did the research use audio or visual recording to collect the data? | Yes, both audio and video were recorded. Interviewers/facilitators had cameras on to help with establishing rapport. Participants had the option to have their cameras on or off, but most chose to keep them on. |
| Field notes | Were field notes made during and/or after the interview or focus group? | Yes |
| Duration | What was the duration of the interviews or focus groups? | Interviews: 45-60 minutes  Focus groups: 90 minutes |
| Data saturation | Was data saturation discussed? | Yes |
| Transcripts returned | Were transcripts returned to participants for comment and/or correction? | No |
| **Domain 3: analysis and findings** | | |
| Data analysis | | |
| Number of data coders | How many data coders coded the data? | Two (TS, JR) – both researchers coded a few focus group transcripts while developing the codebook and both applied codes to a few interview transcripts to confirm consistency; the rest of the transcripts were coded by one researcher with discussions of coding as needed |
| Description of the coding tree | Did authors provide a description of the coding tree? | Five major themes and 18 codes are shown in Table 2 |
| Derivation of themes | Were themes identified in advance or derived from the data? | Themes were derived from the data. Inductive, grounded method of coding. |
| Software | What software, if applicable, was used to manage the data? | Dedoose software (Version 9.0.17, 2021, Los Angeles, CA) |
| Participant checking | Did participants provide feedback on the findings? | No |
| *Reporting* | | |
| Quotations presented | Were participants quotations presented to illustrate the themes/findings? Was each quotation identified? | Yes – quotations in Table 2 identified by role at institution and by whether they were interviewed about experience in quarantine/isolation or participated in a focus group |
| Data and findings consistent | Was there consistency between the data presented and the findings? | Yes |
| Clarity of major themes | Were major themes clearly presented in the findings? | Yes |
| Clarity of minor themes | Is there a description of diverse cases or discussion of minor themes? | Yes |
