## Supplemental File 4 for "Knowledge, attitudes, and practices regarding COVID-19 among university students and employees in Massachusetts, USA: a qualitative study"

**Supplemental file 4:** Description of codebook themes and codes

| Themes and Codes | Description and Examples |
| --- | --- |
| **Theme 1: COVID-19 Knowledge** | |
| Misinformation and Confusion | Inaccurate or incomplete knowledge regarding the science of SARS-CoV-2 or COVID-19 policies, eg. confusion or incorrect knowledge of quarantine or isolation guidelines, contact tracing procedures, or reporting of COVID-19 cases |
| Information Sources | Reported sources of COVID-19-related information, eg. government, news, healthcare providers, friends, family, social media |
| **Theme 2: Stress and Coping** | |
| Direct Stressors from COVID-19 | Stress directly related to COVID-19 infection or fear of infection. Also includes fear of a phone call from contact tracers and grieving for loved ones who have died from COVID-19. |
| Indirect Stressors from COVID-19 | Stress indirectly related to the COVID-19 pandemic, including financial concerns, increased workload, and social isolation. |
| Social Support | Positive experiences or coping mechanisms to deal with stress that involve interpersonal relationships. |
| **Theme 3: Trust** | |
| Science, Government, and Healthcare Trust | Science, government, or healthcare providers mentioned as trusted sources for COVID-19-related information |
| Science, Government, and Healthcare Mistrust | Science, government, or healthcare providers mentioned as being untrustworthy sources for COVID-19-related information |
| Institutional Trust | University of Massachusetts mentioned as a trusted source for COVID-19 related information. Also includes positive comments regarding the university contact tracing program. |
| Institutional Mistrust | University of Massachusetts mentioned as being an untrustworthy source for COVID-19 related information. Also includes negative comments regarding the university contact tracing program, eg. confidentiality concerns for contact tracing, desire to complete quarantine or isolation without entering formal contact tracing process. |
| **Theme 4: Decision-Making** | |
| Testing Choices | Decision-making process for when to obtain a COVID-19 test. Includes behavior changes that occur after negative test results are received, eg. proximity testing with roommates, increased socialization upon receiving negative COVID-19 test results |
| Safer Social Behaviors | Decision-making process for when and how to socialize with others. Includes perceived risk reduction strategies such as gathering outdoors, wearing masks in public, avoiding crowds, and limiting social interactions to a small group of individuals (pod). |
| Negotiation and Peer Pressure | Experiences regarding pressure from others to act in certain ways and navigation of social activities during the pandemic, eg. peer pressure to obtain COVID-19 tests, discussing activity comfort levels with friends |
| Protection of Family Members | Procedures used to protect the health of family members, eg. obtaining a COVID-19 test before visiting family members, refraining from visiting high-risk family members such as grandparents |
| **Theme 5: Institutional Feedback** | |
| Institutional Pride | Positive feedback for the University of Massachusetts Amherst, including favorable comparisons to other institutions. |
| Asymptomatic Testing Pros | Positive feedback for the university asymptomatic testing program, eg. free, short lines, convenient, fast results |
| Asymptomatic Testing Cons | Negative feedback for the university asymptomatic testing program, eg. long lines, required appointments, IT portal problems. |
| Policy Disagreement or Confusion | Confusion regarding university procedures or dislike of university policies, eg. unclear consequences for failing to test regularly, unclear rules for completing wellness checks, dislike of Wellbeing Wednesdays, layoffs, and furloughs |
| Perception of Privileged Groups | Reported perception that certain individuals or groups receive preferential treatment or expanded opportunities during the pandemic, eg. freshman allowed to be on campus, no consequences for those breaking the rules, permission to travel for certain university-sponsored student groups |
